# Adverse effects of remdesivir, hydroxychloroquine, and lopinavir/ritonavir when used for COVID-19: systematic review and meta-analysis of randomized trials

**DOI:** 10.1101/2020.11.16.20232876

**Authors:** Ariel Izcovich, Reed AC Siemieniuk, Jessica J Bartoszko, Long Ge, Dena Zeraatkar, Elena Kum, Assem M. Khamis, Bram Rochwerg, Thomas Agoritsas, Derek K Chu, Shelley L McLeod, Reem A Mustafa, Per O Vandvik, Romina Brignardello-Petersen

## Abstract

**Introduction:** In an attempt to improve outcomes for patients with coronavirus disease 19 (COVID-19), several drugs, such as remdesivir, hydroxychloroquine (with or without azithromycin), and lopinavir/ritonavir, have been evaluated for treatment. While much attention focuses on potential benefits of these drugs, this must be weighed against their adverse effects.

**Methods:** We searched 32 databases in multiple languages from 1 December 2019 to 27 October 2020. We included randomized trials if they compared any of the drugs of interest to placebo or standard care, or against each other. A related world health organization (WHO) guideline panel selected the interventions to address and identified possible adverse effects that might be important to patients. Pairs of reviewers independently extracted data and assessed risk of bias. We analyzed data using a fixed-effects pairwise meta-analysis and assessed the certainty of evidence using the GRADE approach.

**Results:** We included 16 randomized trials which enrolled 8226 patients. Compared to standard care or placebo, low certainty evidence suggests that remdesivir may not have an important effect on acute kidney injury (risk difference [RD] 8 fewer per 1000, 95% confidence interval (CI): 27 fewer to 21 more) or cognitive dysfunction/delirium (RD 3 more per 1000, 95% CI: 12 fewer to 19 more). Low certainty evidence suggests that hydroxychloroquine may increase the risk of serious cardiac toxicity (RD 10 more per 1000, 95% CI: 0 more to 30 more) and cognitive dysfunction/delirium (RD 33 more per 1000, 95% CI: 18 fewer to 84 more), whereas moderate certainty evidence suggests hydroxychloroquine probably increases the risk of diarrhoea (RD 106 more per 1000, 95% CI: 48 more to 175 more) and nausea and/or vomiting (RD 62 more per 1000, 95% CI: 23 more to 110 more) compared to standard care or placebo. Low certainty evidence suggests lopinavir/ritonavir may increase the risk of diarrhoea (RD 168 more per 1000, 95% CI: 58 more to 330 more) and nausea and/or vomiting (RD 160 more per 1000, 95% CI: 100 more to 210 more) compared to standard care or placebo.

**Conclusion:** Hydroxychloroquine probably increases the risk of diarrhoea and nausea and/or vomiting and may increase the risk of cardiac toxicity and cognitive dysfunction/delirium. Remdesivir may have no effect on risk of acute kidney injury or cognitive dysfunction/delirium. Lopinavir/ritonavir may increase the risk of diarrhoea and nausea and/or vomiting. These findings provide important information to support the development of evidence-based management strategies for patients with COVID-19.

## Introduction

As of November 16, 2020, there are 54.6 million cumulative cases of COVID-19 worldwide, and at least 1.3 million deaths.^1^ Several drugs have been used for the treatment of patients with COVID-19, often without high quality evidence demonstrating efficacy. Three drugs that have been used for COVID-19 include remdesivir, hydroxychloroquine with or without azithromycin, and lopinavir/ritonavir. None of these drugs have high certainty evidence evaluating their effectiveness for key patient-important outcomes such as mortality, need for mechanical ventilation, duration of hospital stay or time to clinical improvement.^2^

We are conducting a living systematic review and network meta-analysis to provide a summary of the evidence for all drugs used in the treatment of COVID-19.^2^ Until now, we have not found that any one of these drugs increases the risk of adverse effects leading to drug continuation when compared to standard care or another drug treatment. However, we have not evaluated drug-specific adverse effects, which patients might consider to be important when making decisions about whether to use or not use a drug, particularly in the face of considerable uncertainty regarding their desirable effects.

Building on the work of the living systematic review, the aim of this paper is to summarize the best available evidence addressing drug-specific adverse effects in COVID-19. This evidence synthesis is part of the BMJ-Rapid Recommendations project,^3^ to inform World Health Organization (WHO) Living Guidelines on drugs for treatment of COVID-19.^4,5^

## Methods

We followed the preferred reporting items for systematic reviews and meta-analyses (PRISMA) guidelines for reporting.^6^

### Eligibility criteria

As selected by the linked guideline panel we included randomised clinical trials (RCTs) that included people with suspected, probable, or confirmed COVID-19 comparing remdesivir, hydroxychloroquine, and lopinavir/ritonavir, alone or in combination with other drugs, for treatment against one another or against no intervention, placebo, or standard care, and reported on drug-specific adverse events of interest (see outcome identification below). We included trials regardless of publication status (peer reviewed, in press, or preprint) or language. No restrictions were applied based on severity of COVID-19 illness or setting in which the trial was conducted (outpatient, hospital, ICU, etc). We excluded studies in which remdesivir, hydroxychloroquine, and lopinavir/ritonavir were used for prophylaxis and studies in which different doses of the same intervention were compared.

### Information sources

We performed daily searches from Monday to Friday using the WHO COVID-19 database for eligible studies, which is a comprehensive multilingual source of global literature on COVID-19.^7^ Prior to its merge with the WHO COVID-19 database on 9 October 2020, we also performed daily searches for eligible studies from Monday to Friday in the US Centers for Disease Control and Prevention (CDC) COVID-19 Research Articles Downloadable Database.^8^ To identify RCTs, we filtered the results from the CDC’s database through a validated and highly sensitive machine learning model.^9^ In addition, we searched six Chinese databases. We adapted the search terms for COVID-19 developed by the CDC to the Chinese language. For the Chinese literature search, we also included search terms for randomised trials.

We also used living evidence retrieval services to identify any trials that might have been missed with traditional search methods. These included the Living Overview of the Evidence (L-OVE) COVID-19 Repository by the Epistemonikos Foundation^10^ and the Systematic and Living Map on COVID-19 Evidence by the Norwegian Institute of Public Health, in collaboration with the Cochrane Canada Centre at McMaster University.^11^ We searched all English information sources from 1 December 2019 to 27 October 2020, and the Chinese literature from inception of the databases to 16 October 2020. A complete list of information sources is available in supplementary text 1.

### Study selection

Using systematic review software, Covidence,^12^ following training and calibration exercises, pairs of reviewers independently screened all titles and abstracts, followed by full texts of trials that were identified as potentially eligible. A third reviewer adjudicated conflicts.

### Data collection

For each eligible trial, pairs of reviewers extracted data independently using a standardised, pilot-tested data extraction form. Reviewers collected information on trial characteristics (trial registration, publication status, study status, design), participant characteristics (country, age, sex, smoking habits, comorbidities), and outcomes of interest. Reviewers resolved discrepancies by discussion and, when necessary, with adjudication by a third party.

#### Outcome identification

A linked WHO-BMJ Rapid Recommendations guideline panel^4^ consisting of patients, clinicians, and research methodologists with representation from all WHO geographic regions provided input on potentially important adverse effects of the medications. If any of the panelists believed a specific adverse effect was possible and might influence the decision to use or not use each drug, it was included in this systematic review as an outcome of interest. Panelists were asked to focus on adverse effects important to patients, rather than surrogate measures. For example, we considered clinically important cardiac toxicity including arrhythmias important, but did not consider changes to the QT interval important.

The panel identified specific adverse effects for each drug. For remdesivir, we included acute kidney injury. For hydroxychloroquine and hydroxychloroquine with azithromycin, we included cardiac toxicity, diarrhoea, and nausea and/or vomiting. For lopinavir/ritonavir, we included acute kidney injury, diarrhoea, and nausea and/or vomiting. For all of the drugs, we included cognitive dysfunction/delirium and fatigue. We included studies in which researchers used any definitions of these outcomes. In cases in which the definitions did appropriately reflect what is important to patients, we rated down the certainty of the evidence for indirectness (see certainty of the evidence below).

### Risk of bias within individual studies

For each eligible trial and outcome, following training and calibration exercises, reviewers used a revision of the Cochrane tool for assessing risk of bias in RCTs (RoB 2.0)^13^ to rate trials as either at i) low risk of bias, ii) some concerns—probably low risk of bias, iii) some concerns— probably high risk of bias, or iv) high risk of bias, across the following domains: bias arising from the randomisation process; bias due to departures from the intended intervention; bias due to missing outcome data; bias in measurement of the outcome; bias in selection of the reported results, including deviations from the registered protocol; and bias arising from early termination for benefit. We rated trials at high risk of bias overall if one or more domains were rated as “some concerns—probably high risk of bias” or as “high risk of bias”, and as low risk of bias overall if all domains were rated as “some concerns—probably low risk of bias” or “low risk of bias”. Reviewers resolved discrepancies by discussion and, when not possible, with adjudication by a third party.

### Data synthesis

#### Measures of effect and statistical analysis

We summarised the effect of interventions on dichotomous outcomes using odds ratios (ORs) and corresponding 95% confidence intervals (CIs). We conducted frequentist fixed-effects pairwise meta-analyses using the R package “meta” in RStudio Version 1.3.1093.^14^ We used fixed rather than random effects for the primary analysis because for many of the interventions, the evidence consisted of two or fewer trials or there were several studies with few events. For outcomes in which there were more than one trial with no events in both groups, we meta-analysed the data using risk differences (RD) rather than odds ratios.

### Certainty of the evidence

We assessed the certainty of evidence using the grading of recommendations assessment, development and evaluation (GRADE) approach.^15^ Two methodologists with experience in using GRADE rated each domain for each comparison separately and resolved discrepancies by consensus. We rated the certainty for each comparison and outcome as high, moderate, low, or very low, based on considerations of risk of bias, inconsistency, indirectness, publication bias, and imprecision. We made judgments of imprecision using a minimally contextualised approach with the null effect as a threshold. This minimally contextualised approach considers whether the CI includes the null effect, or, when the point estimate is close to the null effect, whether the CI lies within the boundaries of small but important benefit and harm.^16^ Additionally we analysed if the total number of patients included in the meta-analysis was less than the required number of patients generated by a conventional sample size calculation for a single adequately powered trial to define if optimal information size (OIS) was met. For some of the interventions, extensively implemented in other clinical scenarios, we used indirect evidence to complement the certainty of evidence judgments. We created GRADE evidence summaries (Summary of Findings tables) using the MAGIC Authoring and Publication Platform (www.magicapp.org) to provide user friendly formats for clinicians and patients and to allow re-use in the context of clinical practice guidelines for COVID-19.^4,5^ We calculated the absolute risks and risk differences from the ORs (and their CIs) and the mean risk in the control groups across all of the included trials. In cases where no events were reported in the control arm of any of the included studies, we calculated baseline risks based on other comparisons for the same outcome.

### Subgroup and sensitivity analyses

We performed Bayesian random-effects meta-analysis using the *bayesmeta* package.^17^ We used a plausible prior for the variance parameter and a uniform prior for the effect parameter, as suggested in an empirical study using pre-specified empiric priors as a sensitivity analysis for all comparisons.^18^ We did not conduct any subgroup analyses.

## Results

### Study identification

After screening 14,806 titles and abstracts and 300 full texts, we included sixteen unique RCTs with 8,152 patients that informed on drug-specific adverse events (figure 1).^19-34^ We did not identify any additional eligible RCTs through the living evidence retrieval services. Two studies reported adverse effects for remdesivir,^19,33^ ten for hydroxychloroquine,^21-26,29-32^ one for hydroxychloroquine plus azithromycin,^21^ and four for lopinavir/ritonavir.^20,27,28,34^ Of the sixteen eligible RCTs, 13 have been published in peer reviewed journals, and 3 only as preprints.^22,24,26^ All of the trials were registered, published in English and most evaluated treatment in patients admitted to hospital with COVID-19 (15/16; 93.7%). Most of the trials were conducted in China (10/16; 62.5%). Table 1 presents the characteristics of the included studies. Additional study characteristics, outcome data, and risk of bias assessments for each study are available in the supplementary file.

**Table 1.**
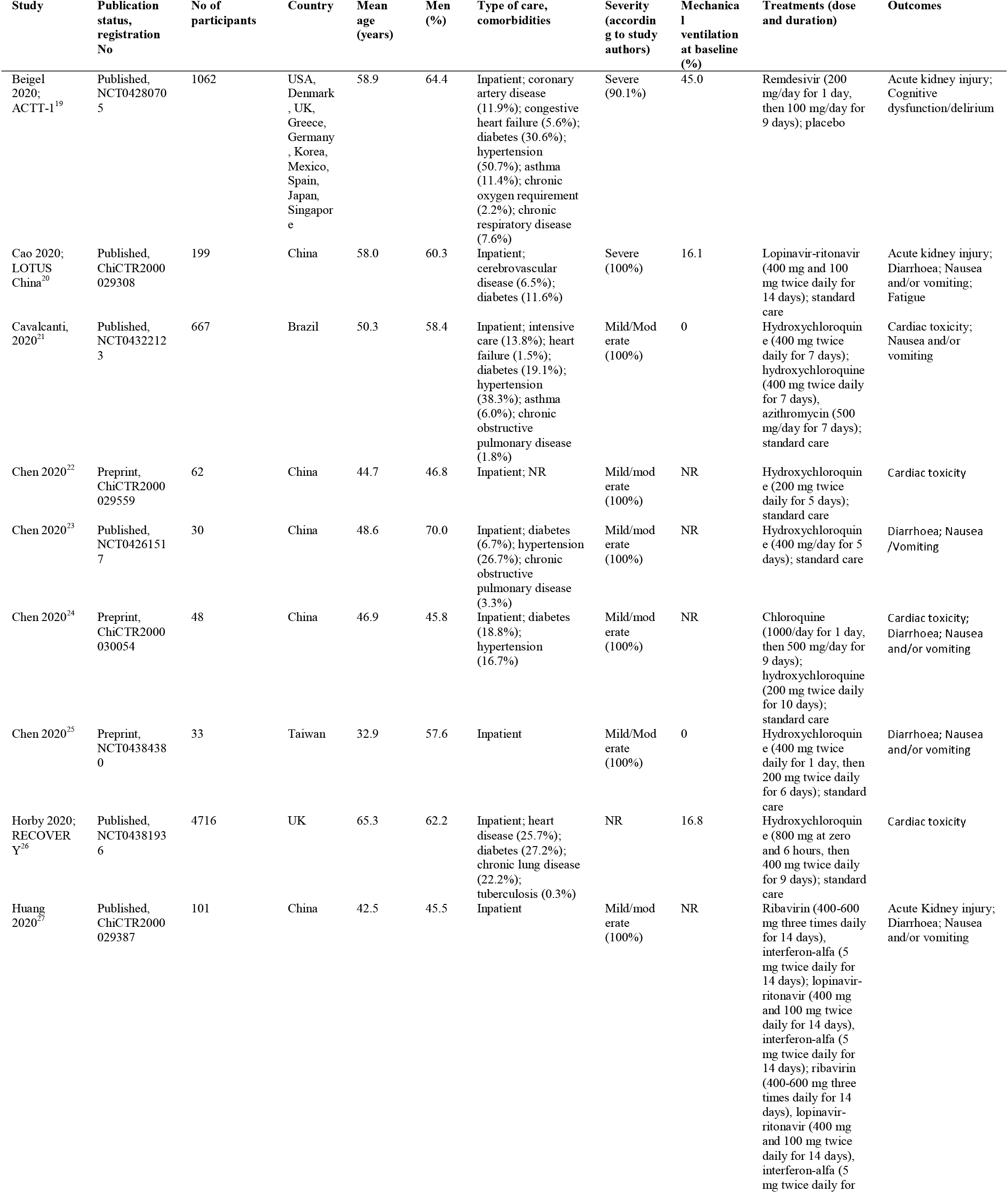
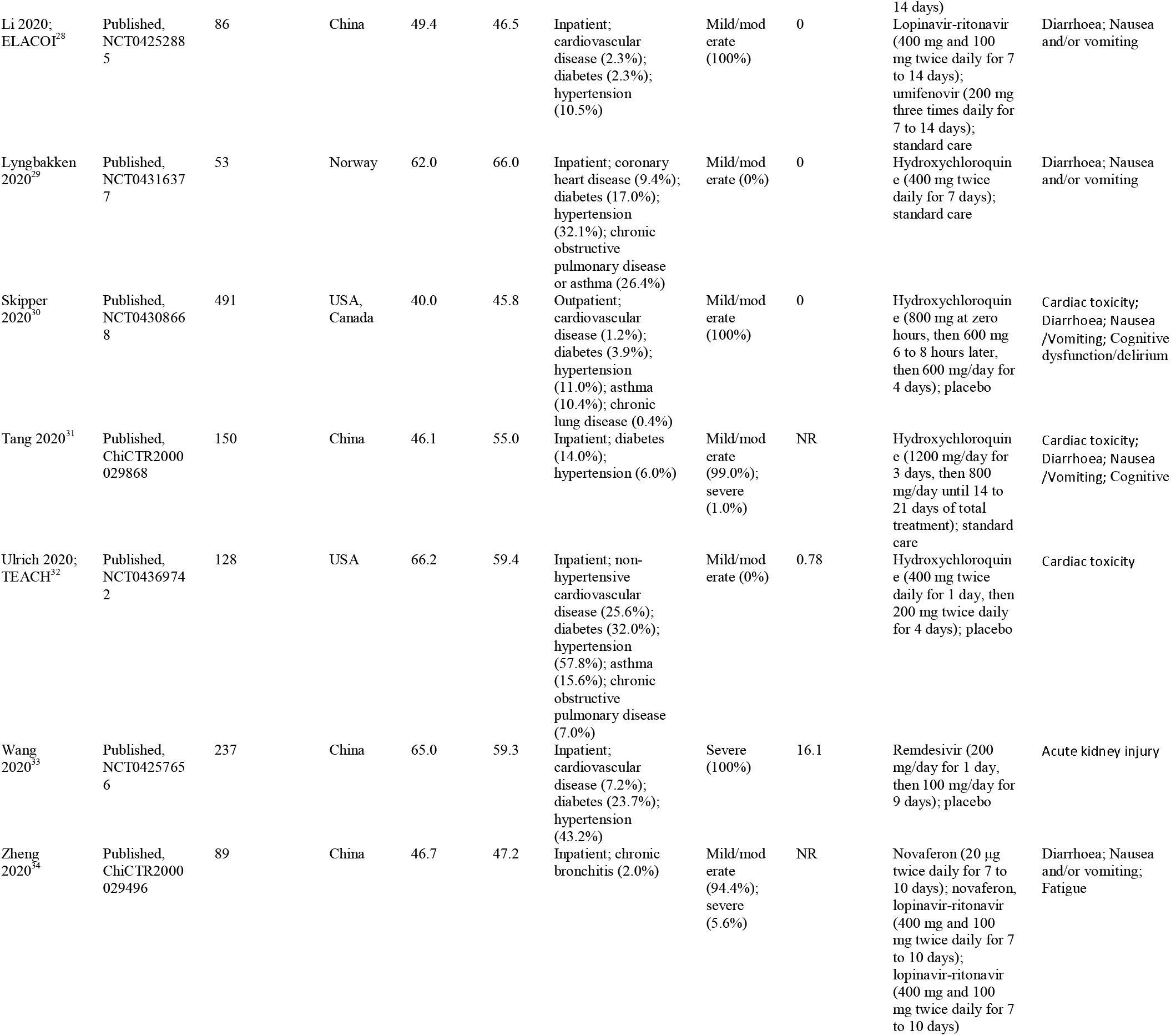
Characteristics of the included trials.

**Figure 1.**
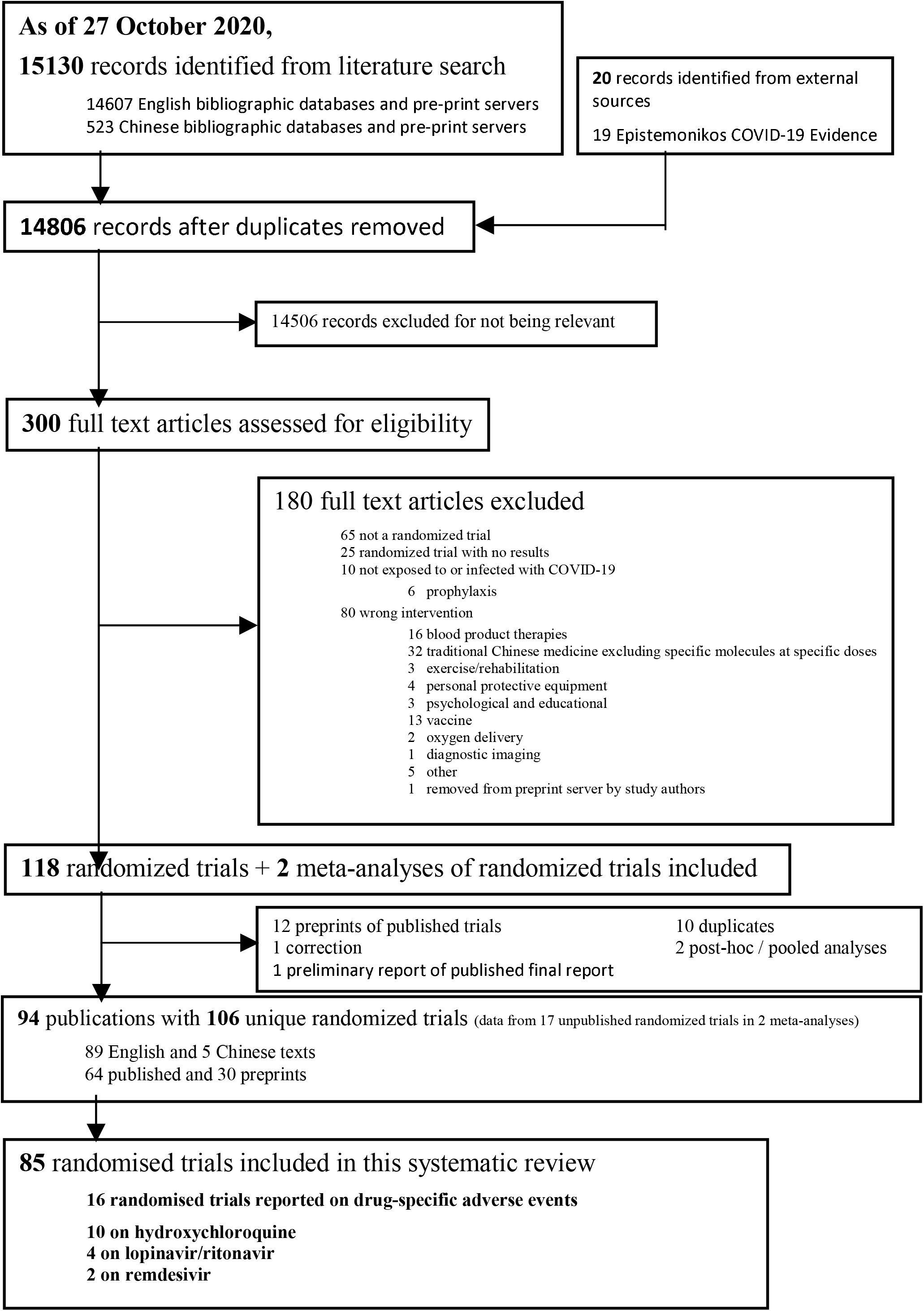
Study selection (PRISMA form LNMA – Jess can help. The bottom cell should have number of trials for each drug separately.)

### Risk of bias in included studies

Supplementary figure 1 presents the risk of bias assessment of the 16 included studies for each outcome. Overall and domain specific risk of bias judgments did not differ between the outcomes reported in each individual study, and most of the studies (13/16, 81.2%) presented significant methodological limitations.

### Adverse effects of the interventions (Table 2.)

#### Remdesivir

Two studies^19,33^ including 1,281 patients reported on remdesivir specific adverse events. Both studies reported on acute kidney injury and one study^19^ including 1,048 patients reported on cognitive dysfunction/delirium. No studies reported on fatigue.

**Table 2.**
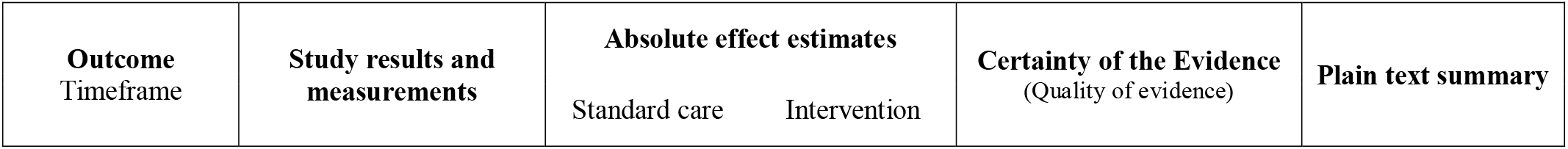
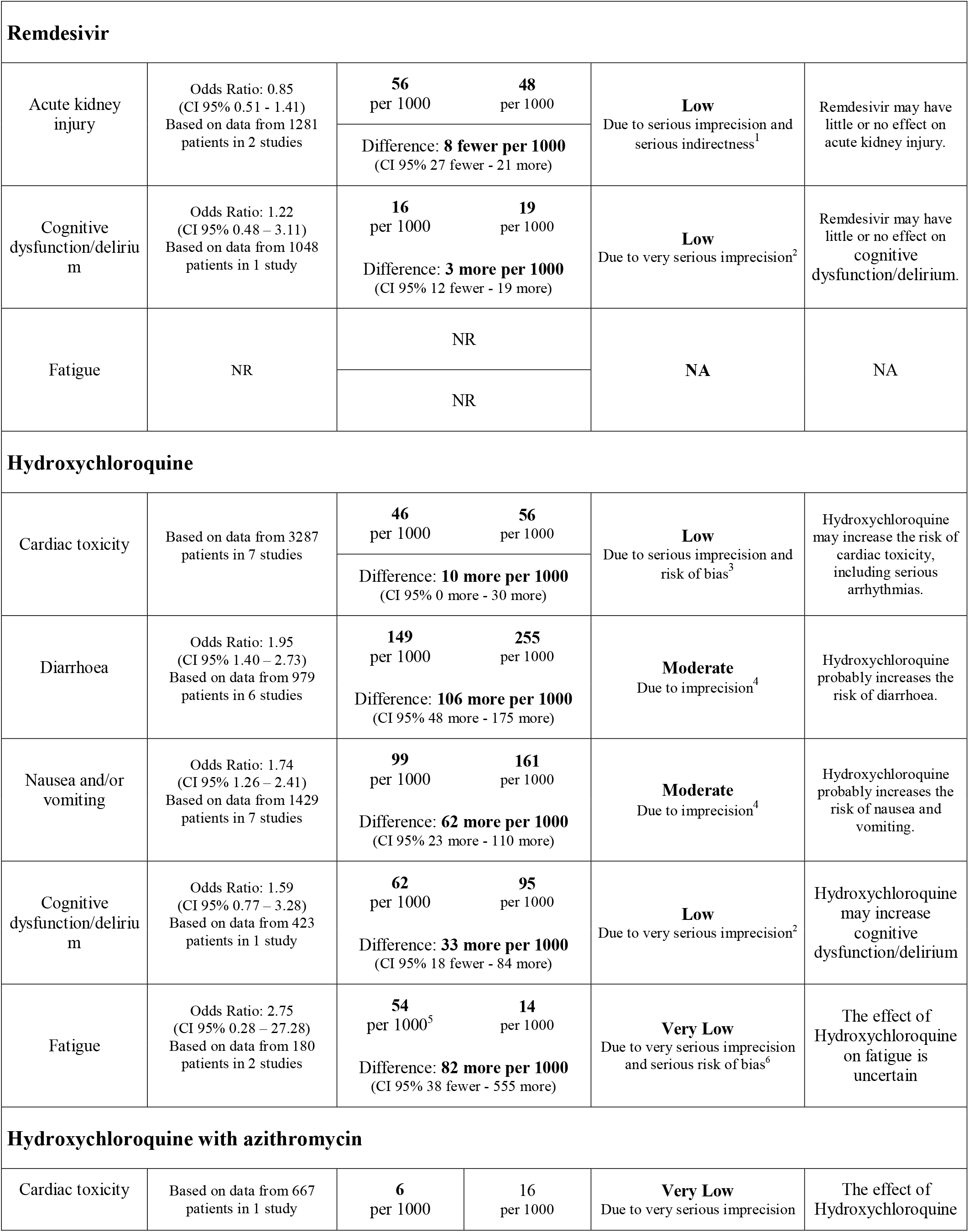
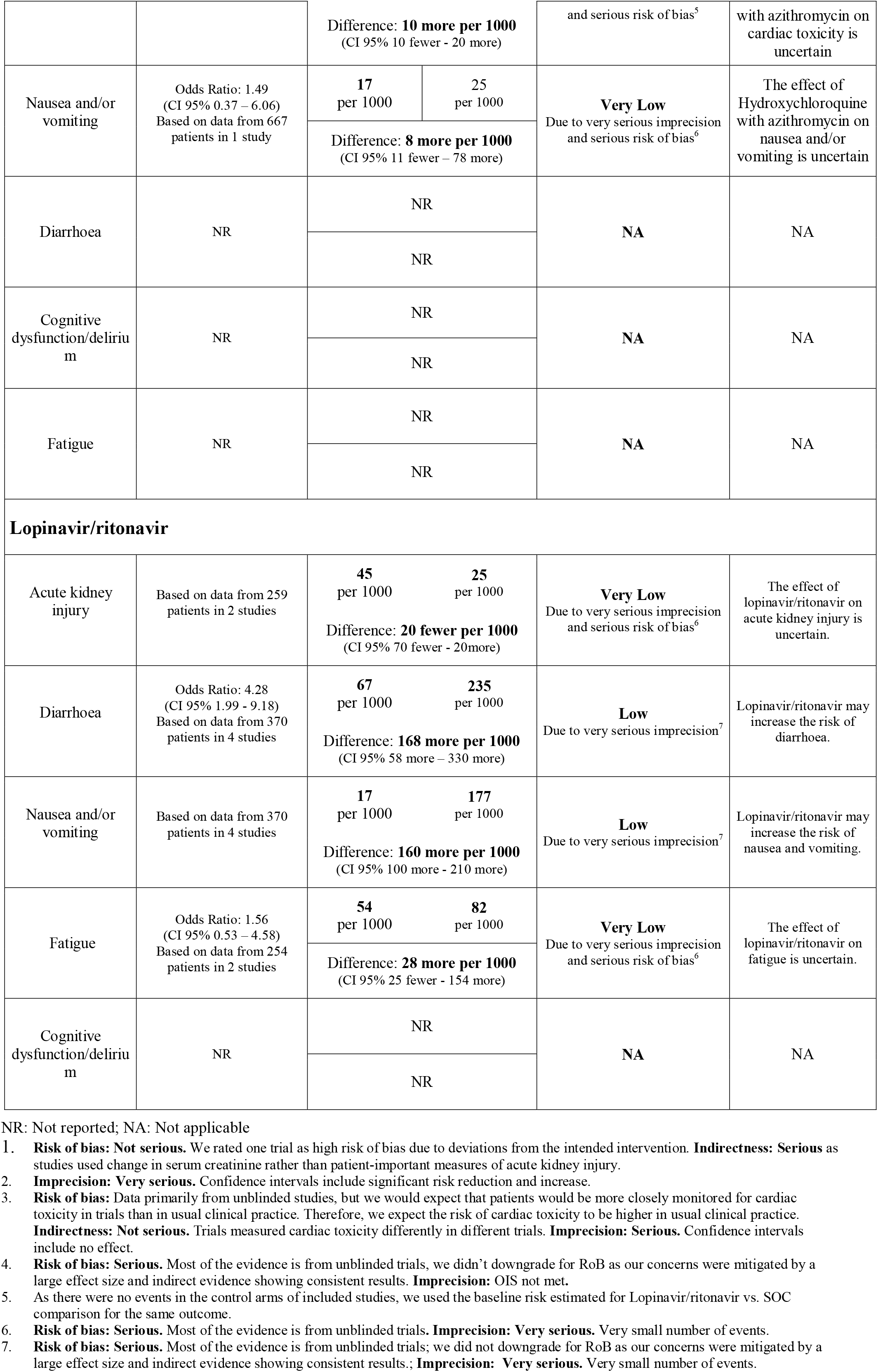
Summary of findings table

##### Acute Kidney Injury

Remdesivir may have little or no effect on acute kidney injury when compared to placebo (OR 0.85, 95% CI: 0.51 to 1.41; RD 8 fewer per 1,000 participants, 95% CI: 27 fewer to 21 more) (Supplementary figure 2.). The certainty of the evidence was low because of serious imprecision and serious indirectness (studies used change in serum creatinine rather than patient-important measures of acute kidney injury like renal replacement therapy requirement).

##### Cognitive dysfunction/delirium

Remdesivir may have little or no effect on cognitive dysfunction/delirium when compared to placebo (OR 1.22, 95% CI: 0.48 to 3.11; RD 3 more per 1,000 participants, 95% CI: 12 fewer to 19 more). The certainty of the evidence was low because of very serious imprecision.

#### Hydroxychloroquine

Ten studies^21-26,29-32^ including 3,663 patients reported on hydroxychloroquine specific adverse events. Seven studies including 3,287 patients reported cardiac toxicity,^21,22,24,27,30-32^ six trials including 979 patients reported diarrhoea,^23-25,29-31^ seven studies including 1,429 patients^23-26,29-31^ reported nausea and/or vomiting, one study^30^ including 423 patients reported on cognitive dysfunction/delirium and two studies^24,31^ including 180 patients reported on fatigue.

##### Cardiac toxicity

Definitions of cardiac toxicity varied between trials: RECOVERY defined the outcome as new major arrhythmias (supraventricular tachycardia, ventricular tachycardia or fibrillation or atrioventricular block requiring intervention),^26^ two studies as new arrhythmias,^21,30^ and one study as new arrhythmias or cardiac arrest.^32^ The remaining studies did not provide details about cardiac toxicity definition. Hydroxychloroquine may increase the risk of cardiac toxicity when compared to standard care or placebo (RD 10 more per 1,000 participants, 95% CI: 0 more to 30 more) (Supplementary figure 3). The certainty of the evidence was low because of serious imprecision and risk of bias (unblinded studies with possible detection bias).

##### Diarrhoea

Hydroxychloroquine probably increases the risk of diarrhoea when compared to standard care or placebo (OR 1.95, 95% CI: 1.40 to 2.73; RD 106 more per 1,000 participants, 95% CI: 48 more to 175 more) (Supplementary figure 4). The certainty of the evidence was moderate because of imprecision as the optimal information size (OIS) was not met. Although most studies presented methodological limitations, we did not rate down for risk of bias (RoB) as our concerns were mitigated by a large effect size and indirect evidence showing consistent results.^35^

##### Nausea and/or vomiting

Hydroxychloroquine probably increases nausea and vomiting (OR 1.74, 95% CI: 1.26 to 2.41; RD 62 more per 1,000 participants, 95% CI: 23 more to 110 more) (Supplementary figure 5). The certainty of the evidence was moderate because of imprecision as OIS was not met. Although most studies presented methodological limitations, we did not rate down for RoB as our concerns were mitigated by a large effect size and indirect evidence showing consistent results.^35^

##### Cognitive dysfunction/delirium

Hydroxychloroquine may increase cognitive dysfunction/delirium when compared to standard care or placebo (OR 1.59, 95% CI: 0.77 to 3.28; RD 33 more per 1,000 participants, 95% CI: 18 fewer to 84 more). The certainty of the evidence was low because of very serious imprecision.

##### Fatigue

The effect of hydroxychloroquine on fatigue is uncertain when compared to standard care or placebo (OR 2.75, 95% CI: 0.28 to 27.28; RD 82 more per 1,000 participants, 95% CI: 38 fewer to 555 more) (Supplementary figure 6). The certainty of the evidence was very low because of very serious imprecision and serious risk of bias.

#### Hydroxychloroquine with azithromycin

Only one study^21^ including 667 patients reported drug-specific adverse effects for hydroxychloroquine with azithromycin. The study compared hydroxychloroquine with azithromycin, hydroxychloroquine alone, and standard care and reported on cardiac toxicity and nausea and/or vomiting. Other outcomes, including diarrhoea, cognitive dysfunction/delirium or fatigue were not reported.

##### Cardiac toxicity

The effect of hydroxychloroquine with azithromycin on cardiac toxicity is uncertain when compared to standard care or placebo (RD 10 more per 1,000 participants, 95% CI: 10 fewer to 20 more), or hydroxychloroquine alone (RD 0 more per 1,000 participants, 95% CI: 20 fewer to 20 more). The certainty of the evidence was very low because of very serious imprecision and serious risk of bias.

##### Nausea and/or vomiting

The effect of hydroxychloroquine with azithromycin on nausea and vomiting in uncertain when compared to standard care or placebo (OR 1.49, 95% CI: 0.37 to 6.06; RD 8 more per 1,000 participants, 95% CI: 11 fewer to 78 more), or hydroxychloroquine alone (OR 0.54, 95% CI: 0.18 to 1.57; RD 20 fewer per 1,000 participants, 95% CI: 37 fewer to 24 more). The certainty of the evidence was very low because of very serious imprecision and serious risk of bias.

#### Lopinavir/ritonavir

Four studies^20,27,28,34^ including 370 patients reported adverse effects of lopinavir/ritonavir. All four studies reported diarrhoea and nausea and/or vomiting. Two studies including 259 patients reported acute kidney injury^20,27^ and two studies including 254 patients reported fatigue.^27,34^ No studies reported on cognitive dysfunction/delirium.

##### Acute Kidney Injury

The effect of lopinavir/ritonavir on acute kidney injury is uncertain when compared to standard care or placebo (20 fewer per 1,000 participants, 95% CI: 70 fewer to 20 more) (Supplementary figure 7). The certainty of the evidence was very low because of very serious imprecision and serious risk of bias.

##### Diarrhoea

Lopinavir/ritonavir may increase the risk of diarrhoea when compared to standard care or placebo (OR 4.28, 95% CI: 1.99 to 9.18; RD 168 more per 1,000 participants, 95% CI: 58 more to 330 more) (Supplementary figure 8). The certainty of the evidence was low because of very serious imprecision. Although most studies presented methodological limitations, we did not rate down for RoB as our concerns were mitigated by a large effect size and indirect evidence showing consistent results.^36^

##### Nausea and/or vomiting

Lopinavir/ritonavir may increase the risk of nausea and/or vomiting when compared to standard care or placebo (RD 160 more per 1,000 participants, 95% CI: 100 more to 210 more) (Supplementary figure 9). The certainty of the evidence was low because of very serious imprecision. Although most studies presented methodologic limitations, we did not rate down for RoB as our concerns were mitigated by a large effect size and indirect evidence showing consistent results.^36^

##### Fatigue

The effect of lopinavir/ritonavir on fatigue is uncertain when compared to standard care or placebo (OR 1.56, 95% CI: 0.53 to 4.58; 28 more per 1,000 participants, 95% CI: 25 fewer to 154 more) (Supplementary figure 10). The certainty of the evidence was very low because of very serious imprecision and serious risk of bias.

#### Sensitivity analyses

Our interpretation of the results did not substantially change when using a Bayesian random effects model rather than frequentist fixed effects or when pooling relative estimates rather than absolute estimates (supplementary figures 11 to 22).

## Discussion

This systematic review and meta-analysis - directly informing the living WHO guideline for COVID-19 therapeutics - provides a comprehensive overview of the evidence for drug-specific adverse effects of interest for three commonly used drugs for treatment of COVID-19. From 40 interventions included in our living network meta-analysis,^2^ we only included studies reporting on drug specific adverse events for remdesivir, hydroxychloroquine, hydroxychloroquine with azithromycin and lopinavir/ritonavir in this review as these drugs are receiving a high degree of interest. None of these interventions may increase the risk of adverse effects leading to discontinuation, however the certainty of the evidence was low for hydroxychloroquine and moderate for remdesivir, while no information was available for hydroxychloroquine with azithromycin, or lopinavir-ritonavir.^2^ In this review we found moderate certainty evidence that hydroxychloroquine increases the risk of diarrhoea and nausea and/or vomiting and low certainty evidence that it increases the risk of cardiac toxicity and cognitive dysfunction/delirium. For lopinavir/ritonavir we found low certainty evidence that it increases the risk of diarrhoea, and nausea and/or vomiting. Based on low or very low certainty evidence, we did not find evidence that remdesivir or lopinavir/ritonavir increase the risk of acute kidney injury or cognitive defunction/delirium.

### Strengths and limitations of this review

The search strategy was comprehensive with explicit eligibility criteria, and no restrictions on language or publication status. To ensure expertise in all areas, the review team is composed of clinical and methods experts who have undergone training and calibration exercises for all stages of the review process. We assessed the certainty of the evidence using the GRADE approach and interpreted the results considering absolute, rather than relative, effects.

We evaluated only a limited number of adverse effects and interventions, as selected by the linked guideline panel. We included an adverse effect if any panel member believed it might be important to patients when deciding whether to use or not to use a drug. However, there may be other patient-important adverse drug effects that were not prespecified by the panel. Further, some may perceive that excluding surrogate outcomes, such as an increase in liver enzymes or electrocardiogram changes may lead to under-appreciation of potential harms, especially for surrogates that are more closely linked on the causal pathway to patient important harms.

So far there is limited evidence for the harms associated with most drugs as adverse effects were only reported by a limited number of studies. For comparisons with sufficient data, the primary limitation of the evidence was lack of blinding, which might introduce bias through differences in cointerventions or outcome assessment between randomisation groups. However, the large magnitude of effects observed resulted in moderate certainty that hydroxychloroquine causes specific adverse events.

These findings are consistent with “The Living Project” (https://covid-nma.com/), which found an increase in any adverse events with hydroxychloroquine (RR 2.16, 95% CI: 1.21 to 3.86) and lopinavir/ritonavir (RR 2.39, 95% CI: 0.21 to 27.57), but not with remdesivir (RR 1.00, 95% CI: 0.87 to 1.15). However, they did not report on specific adverse events. Other systematic reviews found an increase in the risk of diarrhoea and nausea and/or vomiting with lopinavir-ritonavir^37,38^ and hydroxychloroquine,^38-40^ increase in arrhythmias and QTc interval prolongation with hydroxychloroquine alone,^40-42^ or combined with a macrolide,^43,44^ and no significant increase in renal failure with remdesivir.^45^

## Conclusion

Hydroxychloroquine probably increases the risk of diarrhoea and nausea and/or vomiting and may increase the risk of cardiac toxicity and cognitive dysfunction/delirium. Remdesivir may have no effect on risk of acute kidney injury or cognitive dysfunction/delirium.

Lopinavir/ritonavir may increase the risk of diarrhoea and nausea and/or vomiting. These findings provide important information to support the development of evidence-based management strategies for patients with COVID-19.

## Data Availability

Complete data available in the main manuscript and supplementary materials

