## Supplementary methods for "Adverse effects of remdesivir, hydroxychloroquine, and lopinavir/ritonavir when used for COVID-19: systematic review and meta-analysis of randomized trials"

Appendix

Search sources

**WHO covid-19 database:** Medline (Ovid and PubMed), PubMed Central, Embase, CAB Abstracts, Global Health, PsycInfo, Cochrane Library, Scopus, Academic Search Complete, Africa Wide Information, CINAHL, ProQuest Central, SciFinder, the Virtual Health Library, LitCovid, WHO covid-19 website, CDC covid-19 website, Eurosurveillance, China CDC Weekly, Homeland Security Digital Library, ClinicalTrials.gov, bioRxiv (preprints), medRxiv (preprints), chemRxiv (preprints), and SSRN (preprints).

**US Centers for Disease Control and Prevention (CDC) database:** Medline (Ovid and PubMed), PubMed Central, Embase, CAB Abstracts, Global Health, PsycInfo, Cochrane Library, Scopus, Academic Search Complete, Africa Wide Information, CINAHL, ProQuest Central, SciFinder, the Virtual Health Library, LitCovid, WHO covid-19 website, CDC covid-19 website, Eurosurveillance, China CDC Weekly, Homeland Security Digital Library, ClinicalTrials.gov, bioRxiv (preprints), medRxiv (preprints), chemRxiv (preprints), and SSRN (preprints).

**Chinese databases:** Wanfang, Chinese Biomedical Literature, China National Knowledge Infrastructure, VIP, Chinese Medical Journal Net (preprints), and ChinaXiv (preprints).

**Living Overview of the Evidence (L-OVE) COVID-19 Repository by the Epistemonikos Foundation:** Pubmed/medline (updated several times a day), EMBASE (updated weekly), CINAHL (updated weekly), PsycINFO (updated weekly), LILACS (Latin American & Caribbean Health Sciences Literature) (updated weekly), Wanfang Database (updated every 2 weeks), CBM - Chinese Biomedical Literature Database (updated every 2 weeks), CNKI - Chinese National Knowledge Infrastructure (updated every 2 weeks), VIP - Chinese Scientific Journal Database (updated every 2 weeks), IRIS (WHO Institutional Repository for Information Sharing) (updated weekly), IRIS PAHO (PAHO Institutional Repository for Information Sharing)) (updated weekly), IBECS - Índice Bibliográfico Español en Ciencias de la Salud (Spanish Bibliographic Index on Health Sciences) (updated weekly), Microsoft Academic (last searched: Sept 4, 2020), ICTRP Search Portal (updated daily), Clinicaltrials.gov (updated daily), ISRCTN registry (updated daily), Chinese Clinical Trial Registry (updated daily), IRCT - Iranian Registry of Clinical Trials (updated daily), EU Clinical Trials Register: Clinical trials for covid-19 (updated daily), NIPH Clinical Trials Search (Japan) - Japan Primary Registries Network (JPRN) (JapicCTI, JMACCT CTR, jRCT, UMIN CTR) (updated daily, via ICTRP search portal), UMIN-CTR - UMIN Clinical Trials Registry (updated daily, via ICTRP search portal), JRCT - Japan Registry of Clinical Trials (updated daily, via ICTRP search portal), JAPIC Clinical Trials Information (updated daily, via ICTRP search portal), Clinical Research Information Service (CRiS), Republic of Korea (updated daily, via ICTRP search portal), ANZCTR - Australian New Zealand Clinical Trials Registry (updated daily, via ICTRP search portal), ReBec - Brazilian Clinical Trials Registry (updated daily, via ICTRP search portal), CTRI - Clinical Trials Registry - India (updated daily, via ICTRP search portal), RPCEC - Cuban Public Registry of Clinical Trials (updated daily, via ICTRP search portal), DRKS - German Clinical Trials Register (updated daily, via ICTRP search portal), LBCTR - Lebanese Clinical Trials Registry (updated daily, via ICTRP search portal), TCTR - Thai Clinical Trials Registry (updated daily, via ICTRP search portal), NTR - The Netherlands National Trial Register (updated daily, via ICTRP search portal), PACTR - Pan African Clinical Trial Registry (updated daily, via ICTRP search portal), REPEC - Peruvian Clinical Trial Registry (updated daily, via ICTRP search portal), SLCTR - Sri Lanka Clinical Trials Registry (updated daily, via ICTRP search portal), medRxiv (updated several times a day), bioRxiv (updated several times a day), SSRN Preprints (updated several times a day), ChinaXiv (updated every 2 weeks), SciELO Preprints (updated weekly), Research Square (updated daily)
