## Supplementary figures for "Adverse effects of remdesivir, hydroxychloroquine, and lopinavir/ritonavir when used for COVID-19: systematic review and meta-analysis of randomized trials"

Supplementary materials

Risk of bias assessment

Supplementary figure 1. Risk of bias assessment

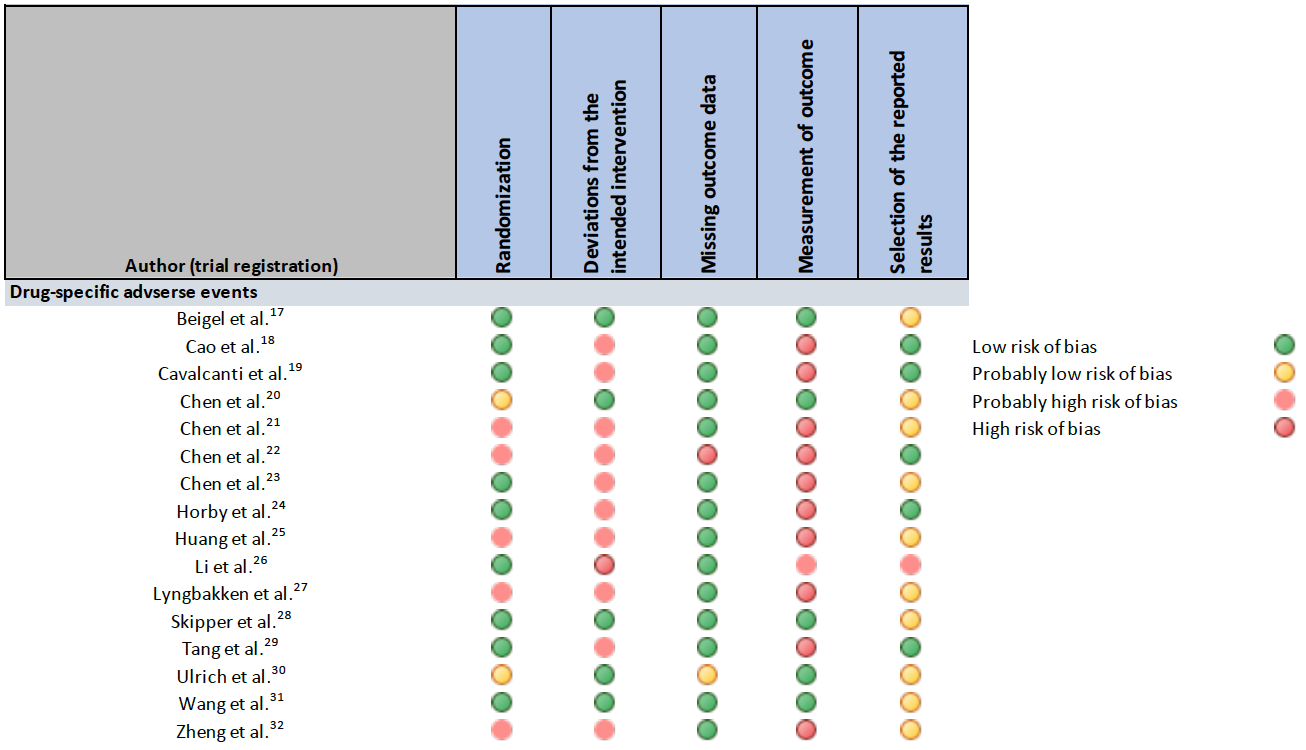

Forest plots: Primary analysis

Supplementary figure 2. Comparison: Remdesivir vs. Standard of care; Outcome: Acute kidney injury

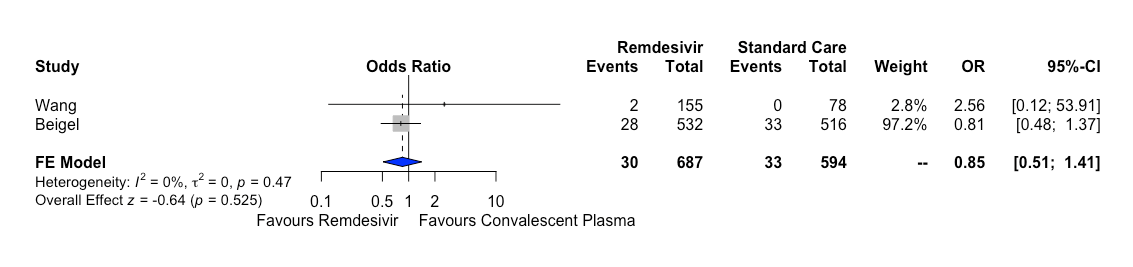

Supplementary figure 3. Comparison: Hydroxychloroquine vs. Standard of care; Outcome: Cardiac toxicity

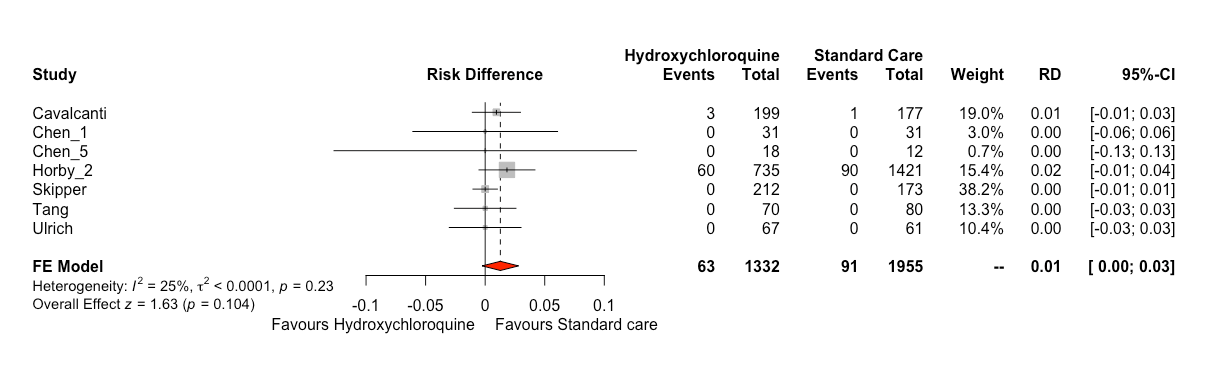

Supplementary figure 4. Comparison: Hydroxychloroquine vs. Standard of care; Outcome: Diarrhoea

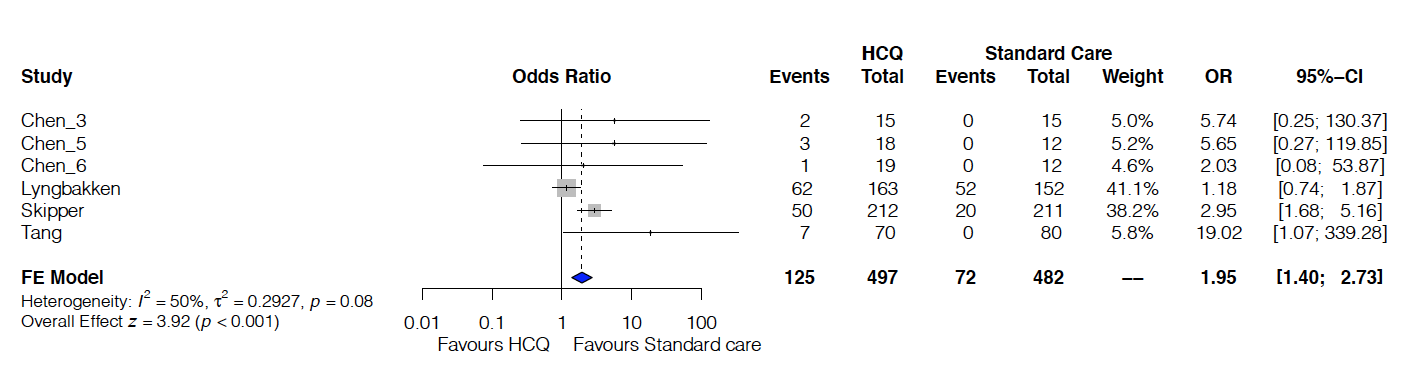

Supplementary figure 5. Comparison: Hydroxychloroquine vs. Standard of care; Outcome: Nausea/Vomiting

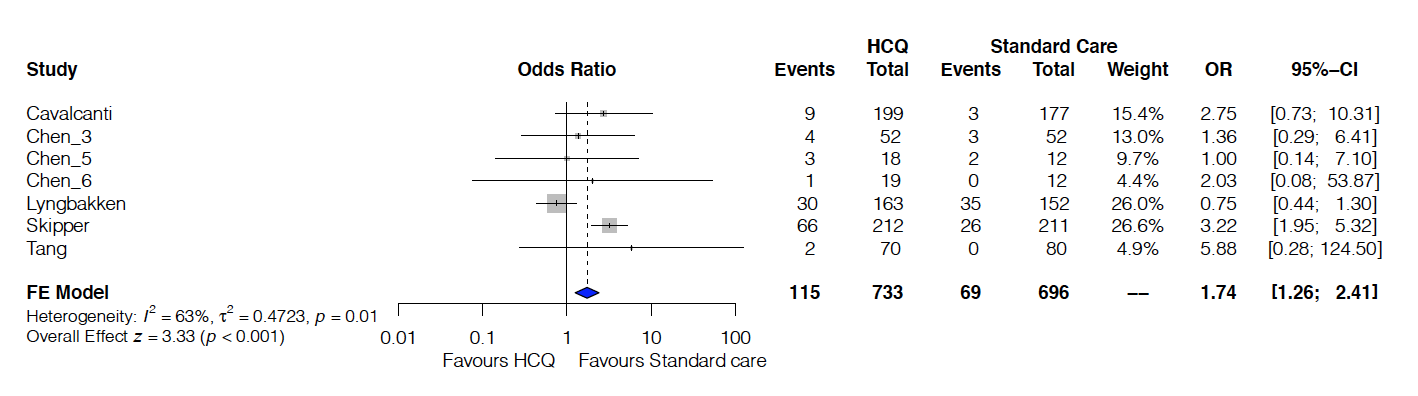

Supplementary figure 6. Comparison: Hydroxychloroquine vs. Standard of care; Outcome: Fatigue

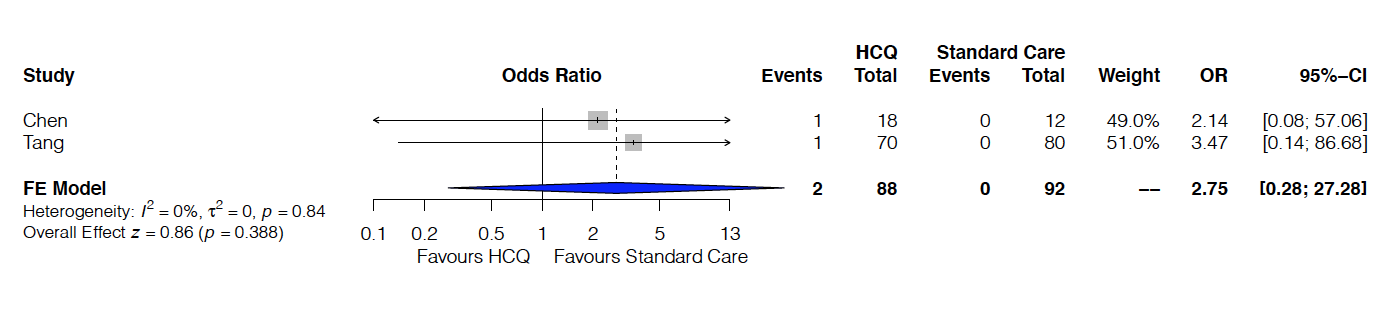

Supplementary figure 7. Comparison: Lopinavir/ritonavir vs. Standard of care; Outcome: Acute Kidney Injury

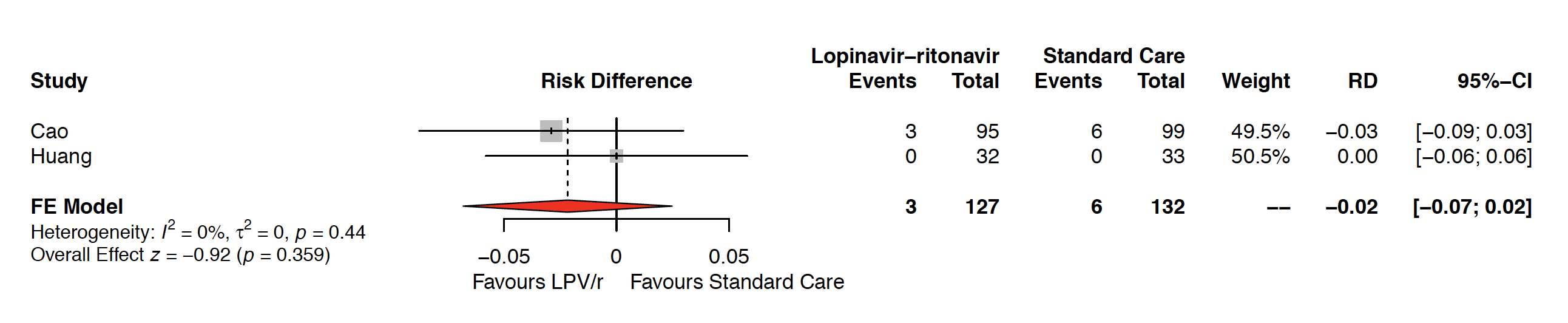

Supplementary figure 8. Comparison: Lopinavir/ritonavir vs. Standard of care; Outcome: Diarrhoea

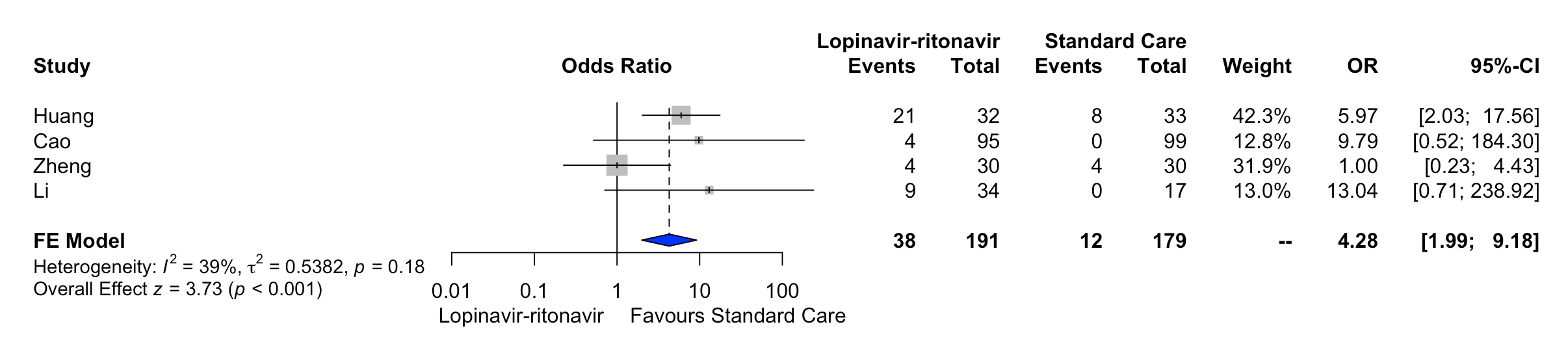

Supplementary figure 9. Comparison: Lopinavir/ritonavir vs. Standard of care; Outcome: Nausea/Vomiting

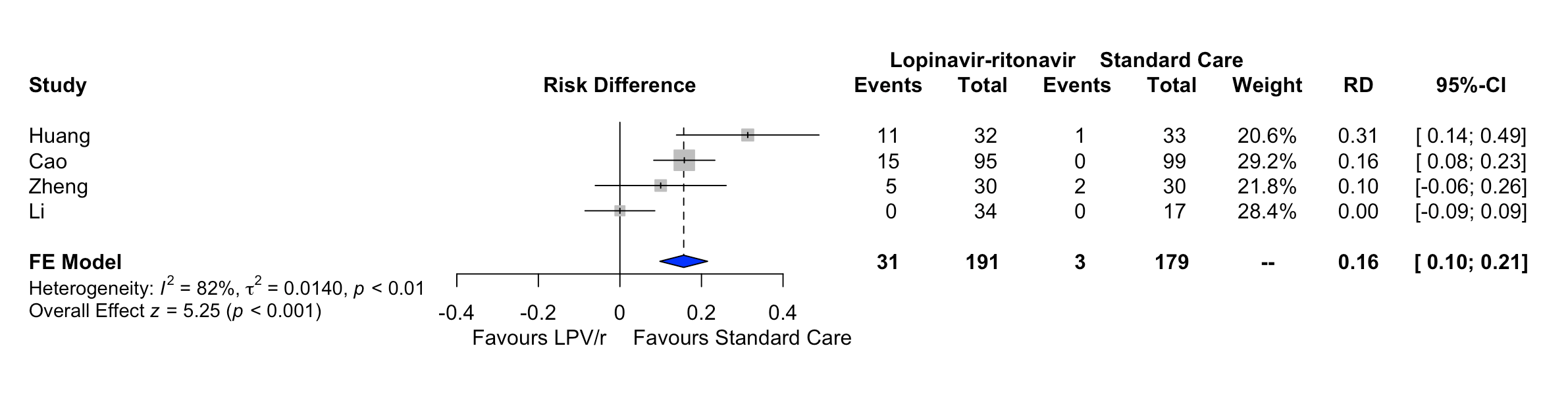

Supplementary figure 10. Comparison: Lopinavir/ritonavir vs. Standard of care; Outcome: Fatigue

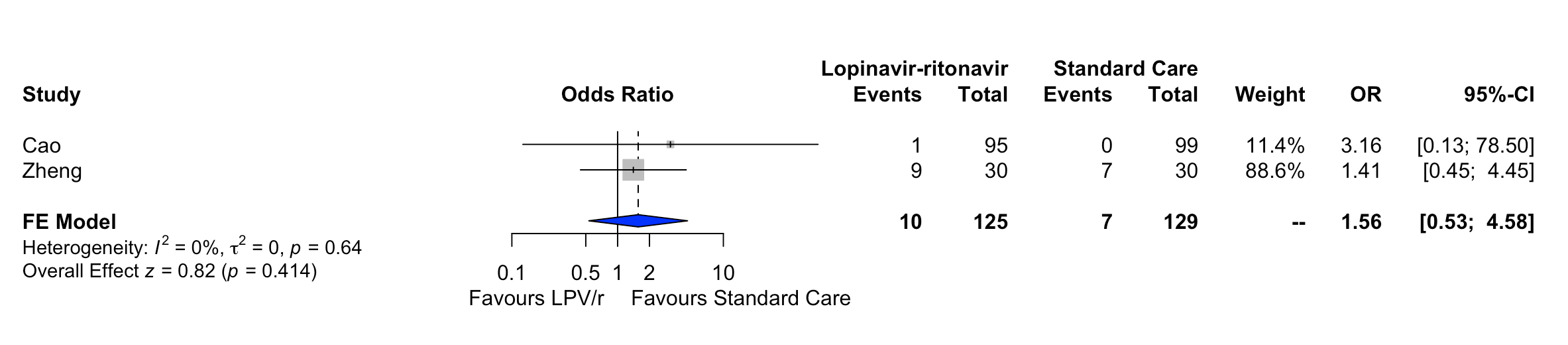

Forest plots: Sensitivity analysis

Supplementary figure 11. Comparison: Lopinavir/ritonavir vs. Standard of care; Outcome: Acute Kidney Injury; Effect estimate: Odds ratio; Analysis: Bayesian meta-analysis

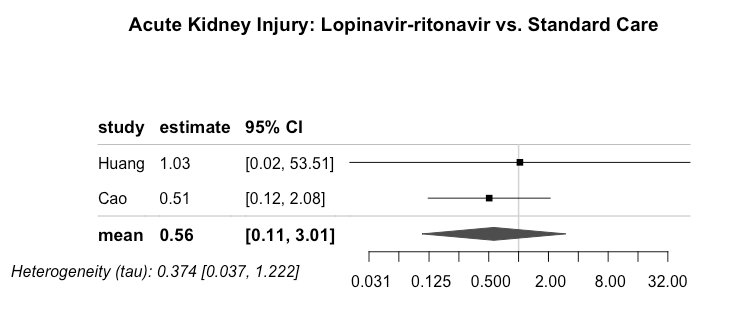

Supplementary figure 12. Comparison: Lopinavir/ritonavir vs. Standard of care; Outcome: Acute Kidney Injury; Effect estimate: Odds ratio, Analysis: Frequentist meta-analysis

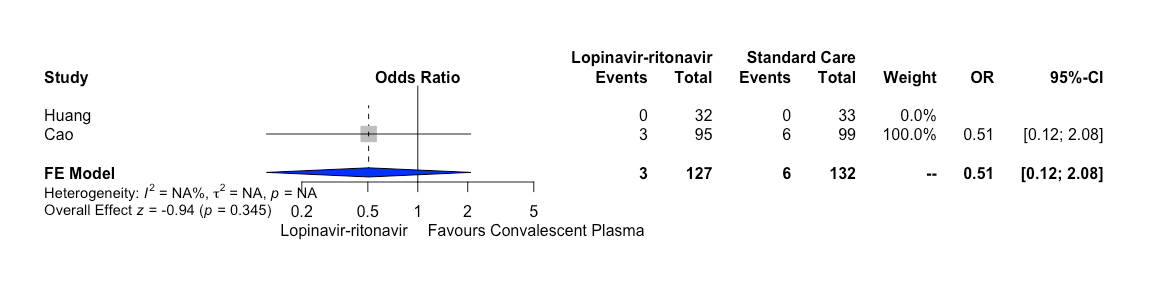

Supplementary figure 13. Comparison: Remdesivir vs. Standard of care; Outcome: Acute Kidney Injury; Effect estimate: Odds ratio, Analysis: Bayesian meta-analysis

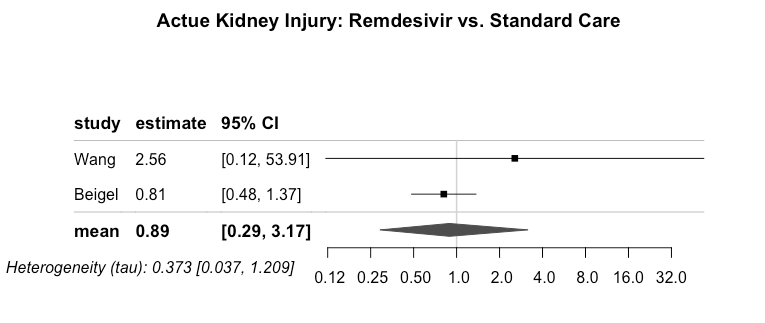

Supplementary figure 14. Comparison: Hydroxicholoroquine vs. Standard of care; Outcome: Cardiac toxicity; Effect estimate: Odds ratio, Analysis: Bayesian meta-analysis

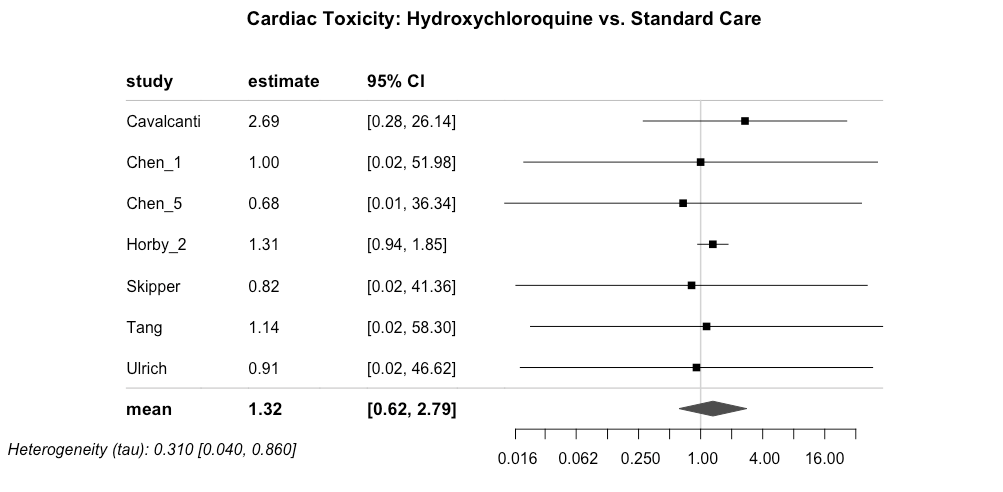

Supplementary figure 15. Comparison: Hydroxicholoroquine vs. Standard of care; Outcome: Cardiac toxicity; Effect estimate: Odds ratio, Analysis: Frecuentist meta-analysis

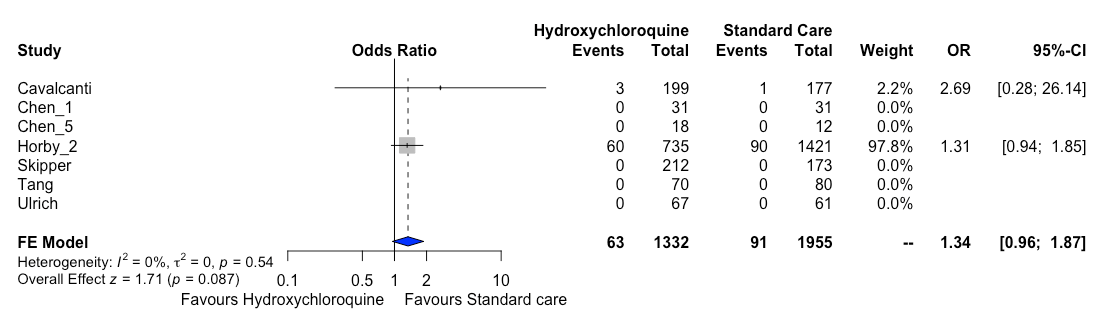

Supplementary figure 16. Comparison: Lopinavir/ritonavir vs. Standard of care; Outcome: Diarrhoea; Effect estimate: Odds ratio; Analysis: Bayesian meta-analysis

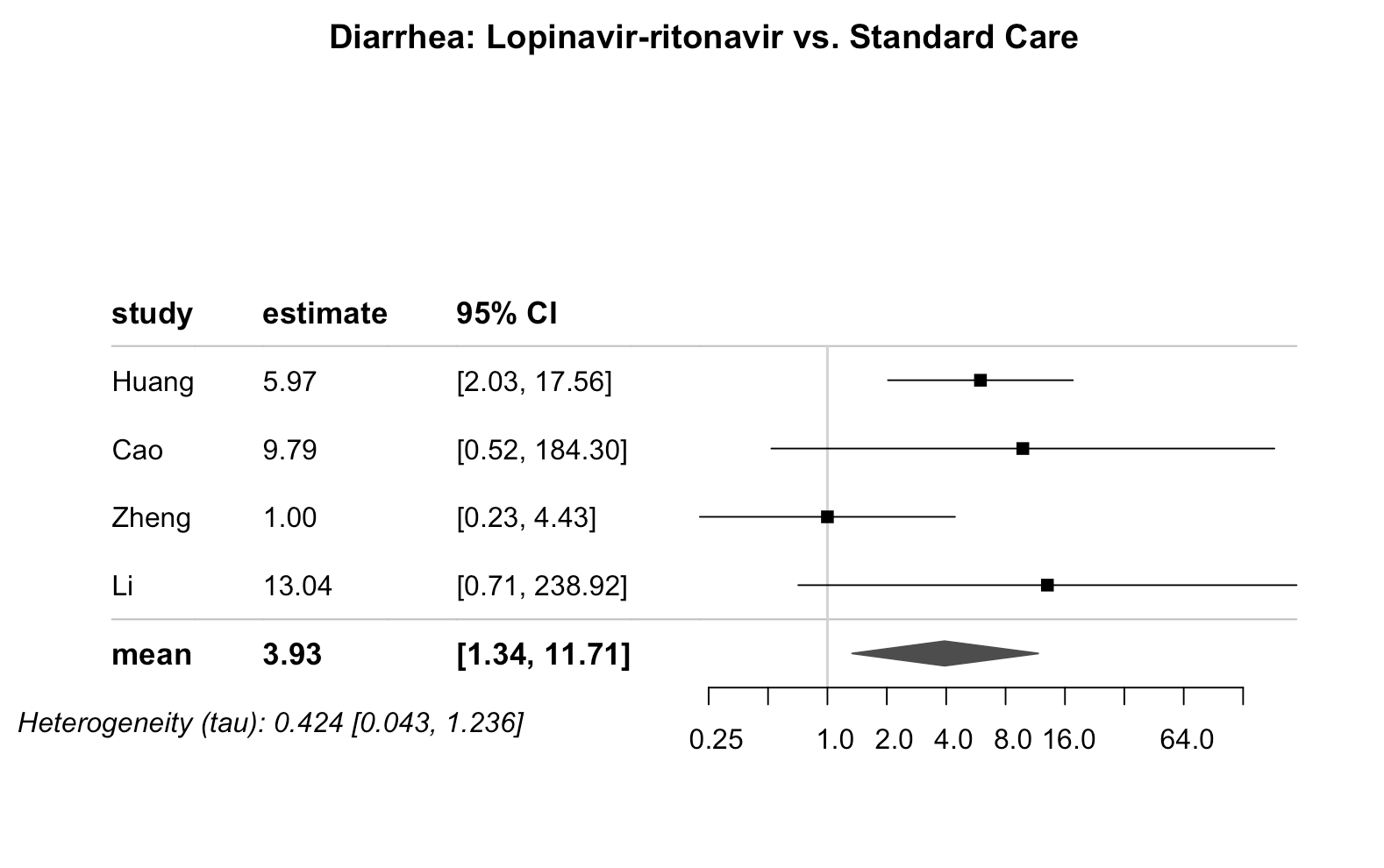

Supplementary figure 17. Comparison: Hydroxychloroquine vs. Standard of care; Outcome: Diarrhoea; Effect estimate: Odds ratio; Analysis: Bayesian meta-analysis

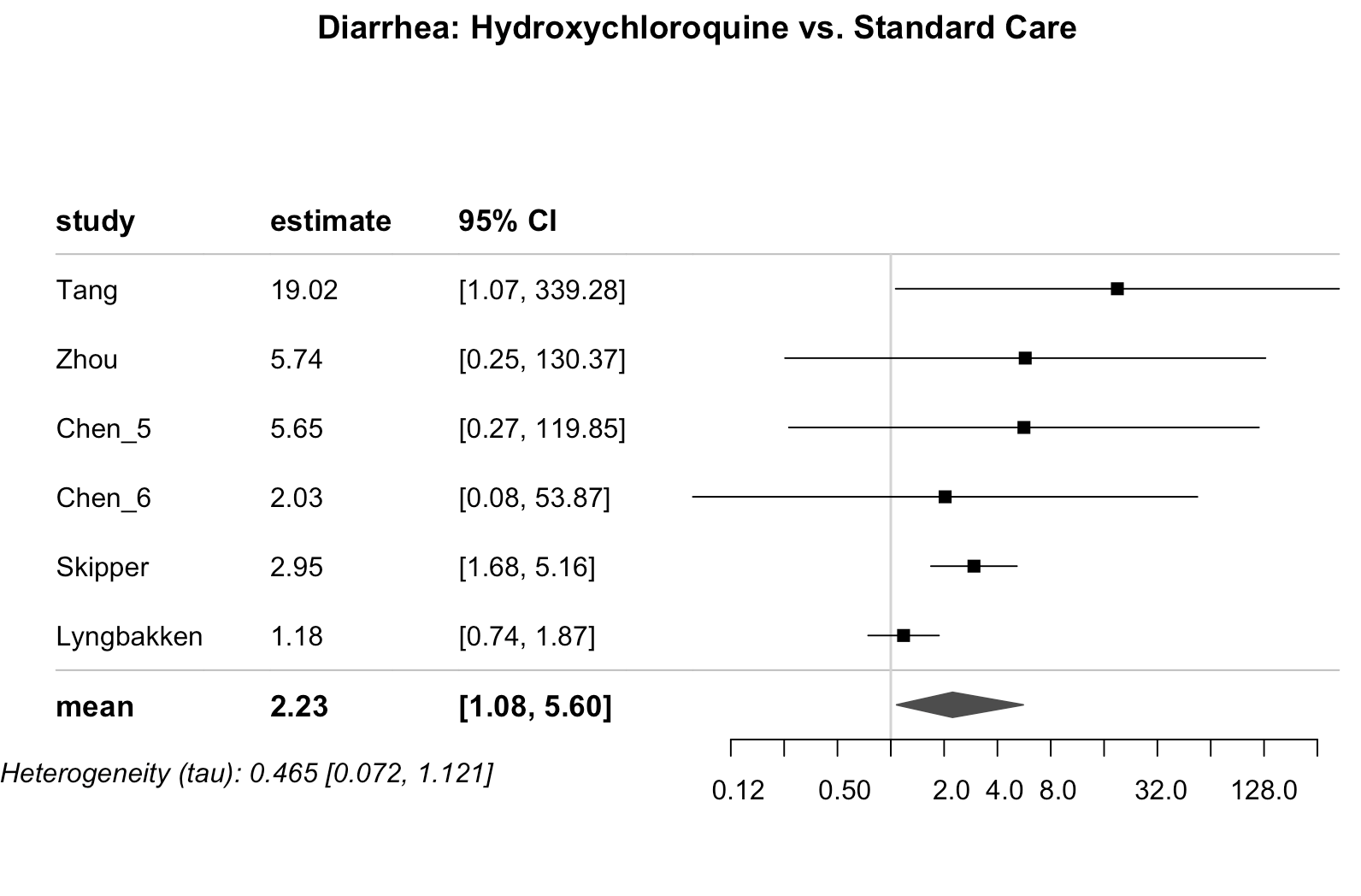

Supplementary figure 18. Comparison: Lopinavir/ritonavir vs. Standard of care; Outcome: Nausea/vomiting; Effect estimate: Odds ratio; Analysis: Bayesian meta-analysis

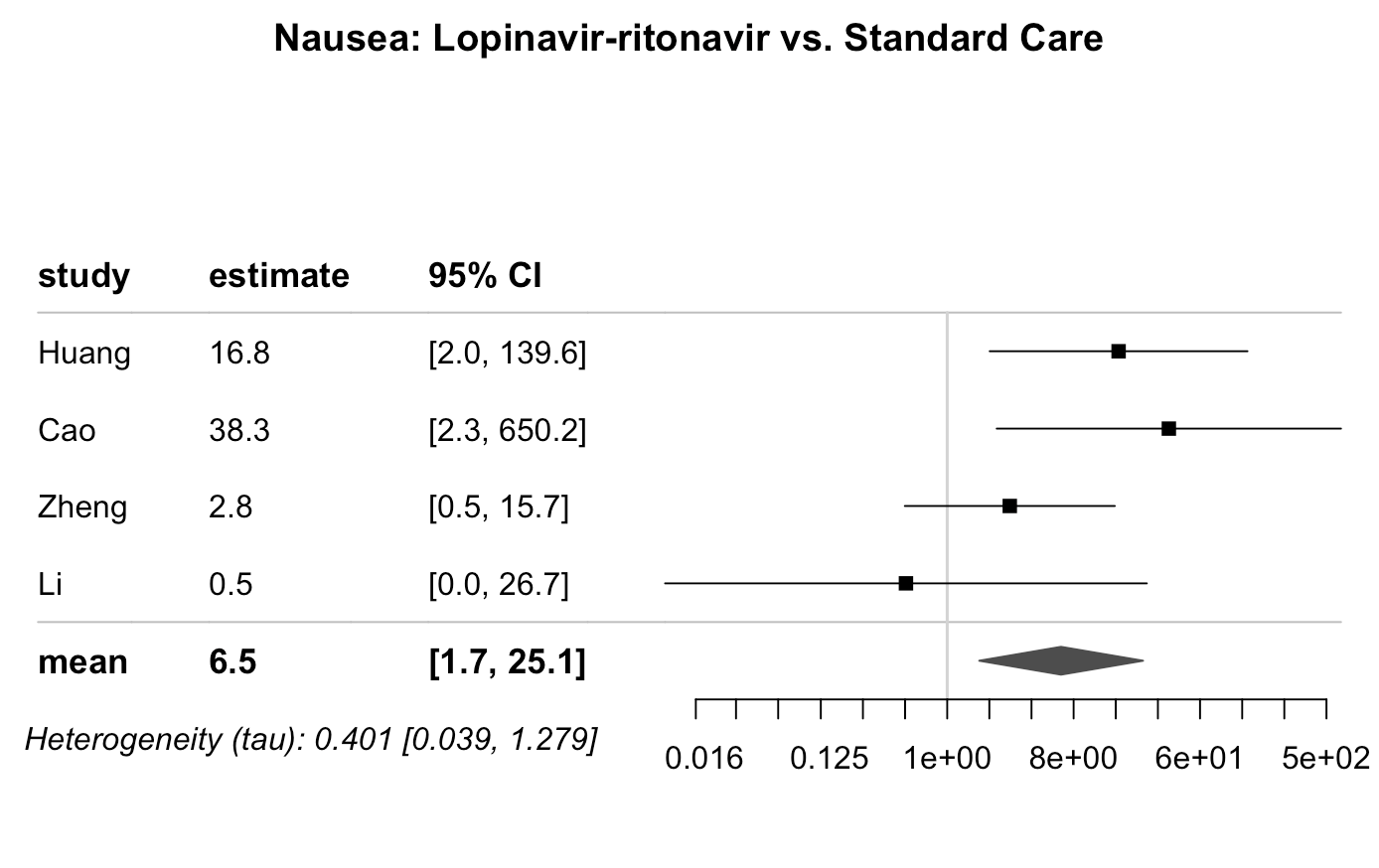

Supplementary figure 19. Comparison: Lopinavir/ritonavir vs. Standard of care; Outcome: Nausea/vomiting; Effect estimate: Odds ratio; Analysis: Frequentist meta-analysis

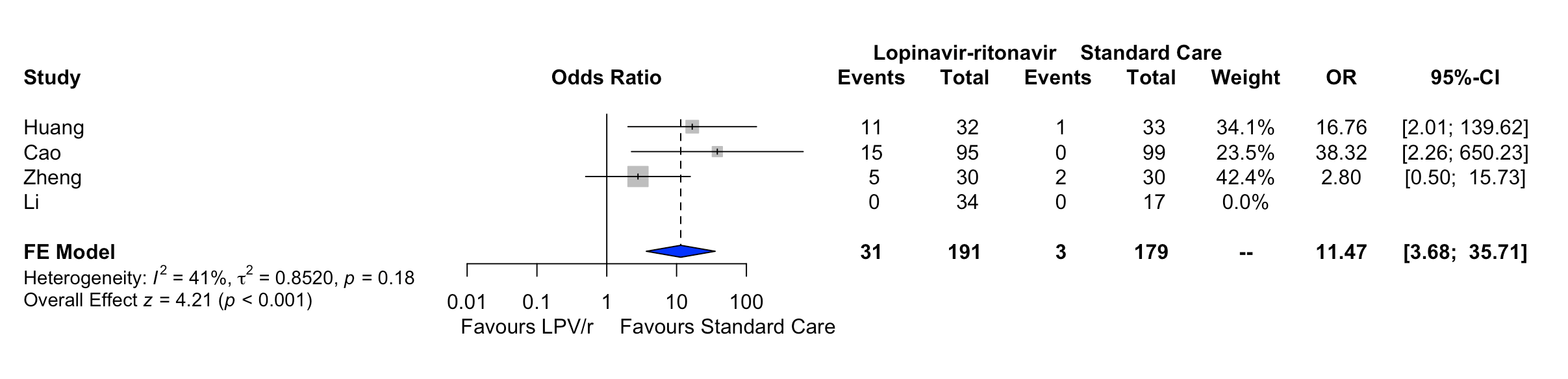

Supplementary figure 20. Comparison: Hydroxychloroquine vs. Standard of care; Outcome: Nausea/vomiting; Effect estimate: Odds ratio; Analysis: Bayesian meta-analysis

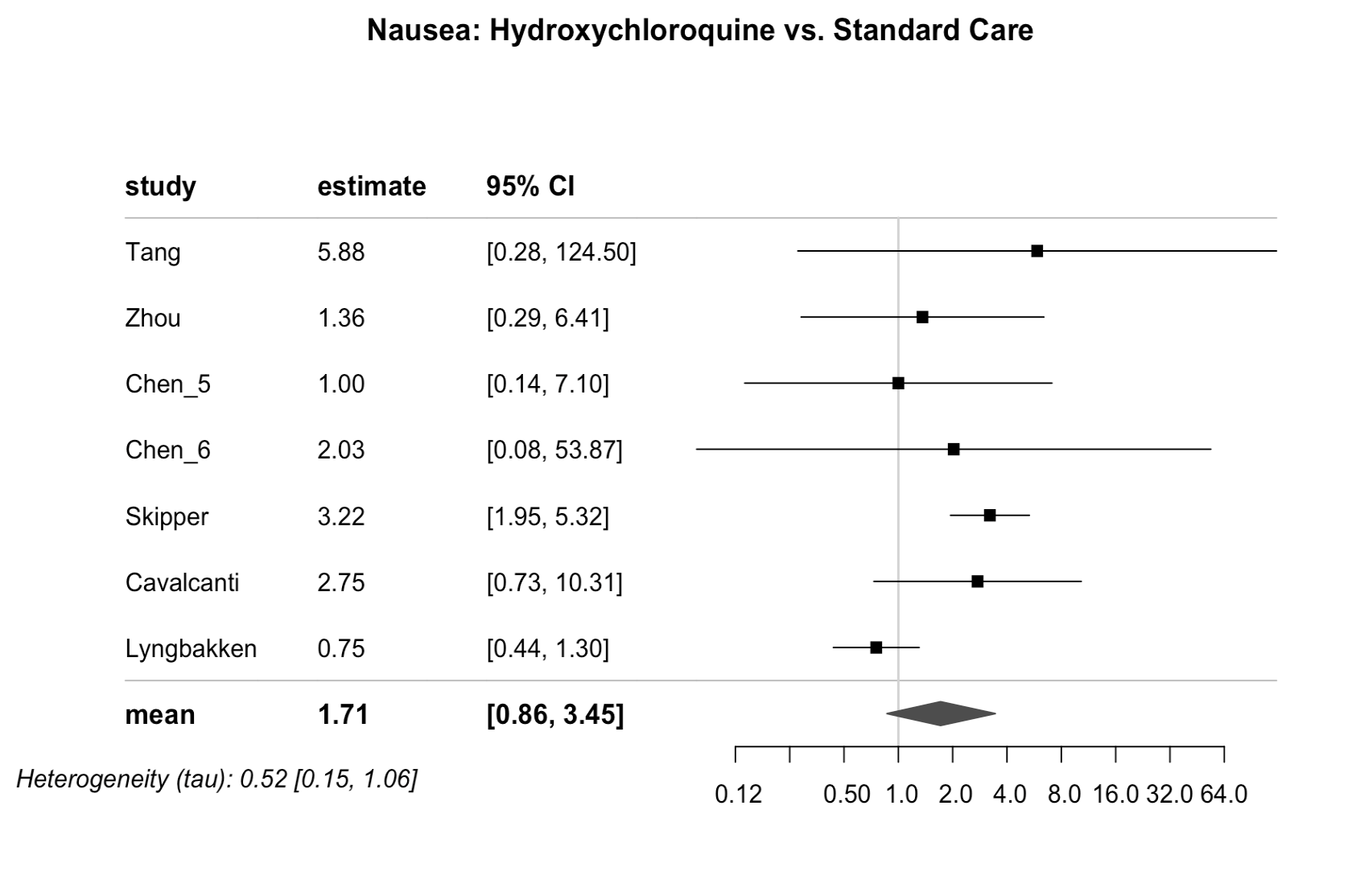

Supplementary figure 21. Comparison: Hydroxychloroquine vs. Standard of care; Outcome: Nausea/vomiting; Effect estimate: Odds ratio; Analysis: Frequentist meta-analysis

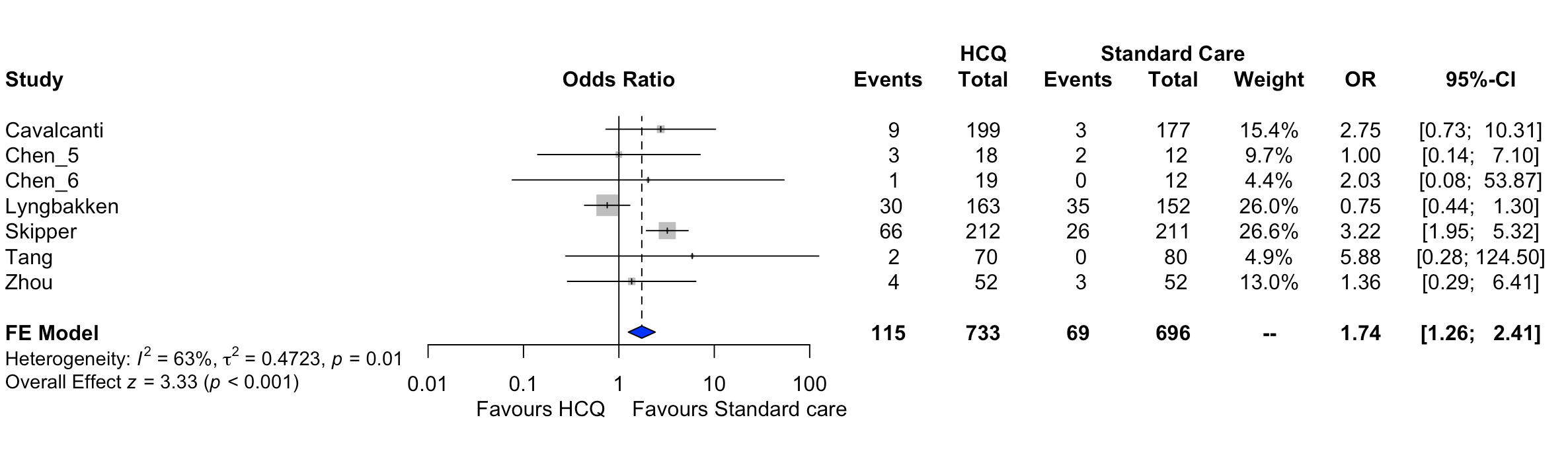

Supplementary figure 22. Comparison: Lopinavir/ritonavir vs. Standard of care; Outcome: Fatigue; Effect estimate: Odds ratio; Analysis: Bayesian meta-analysis

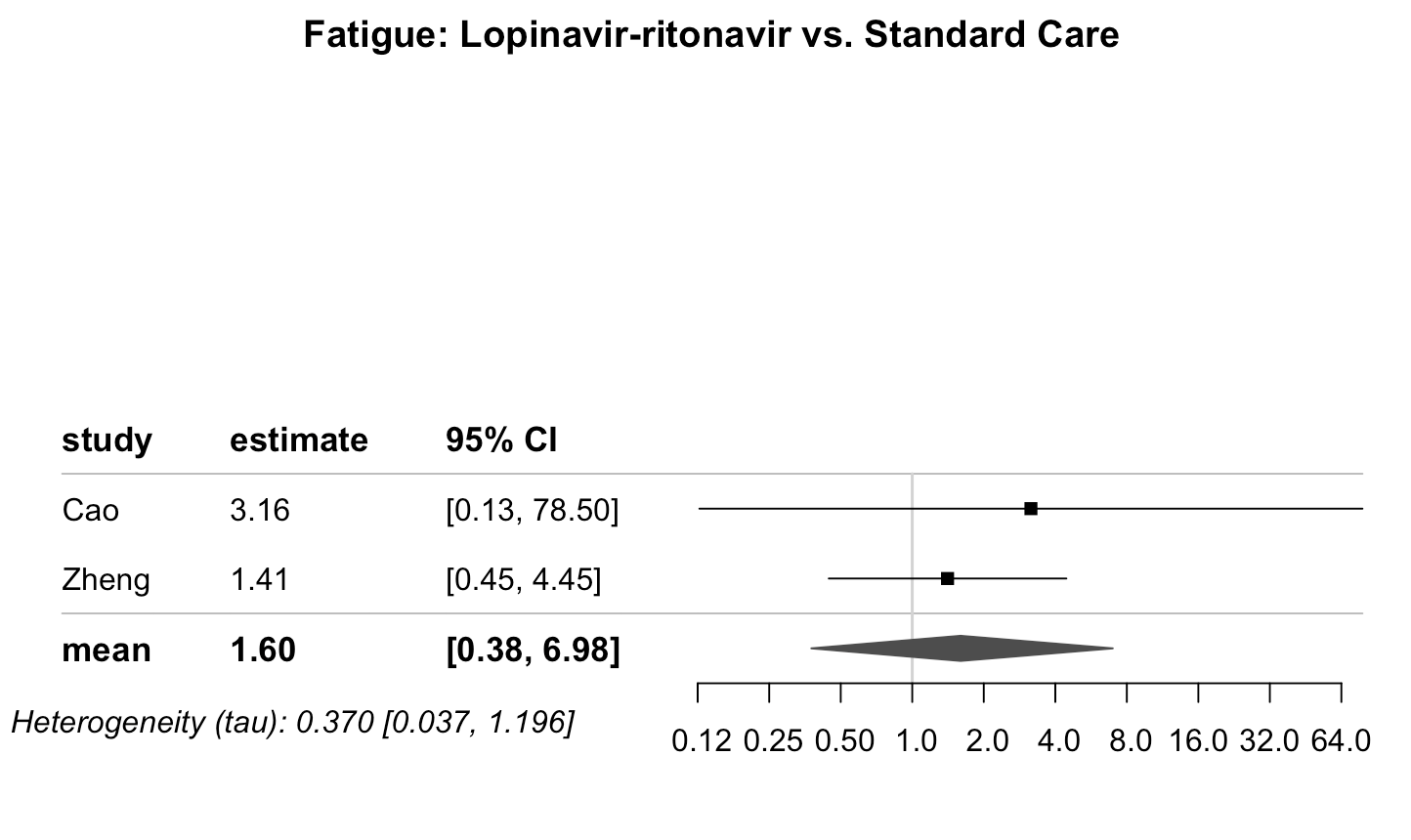
